# FACTORS IMPACTED ON WELLBEING IN ADOLESCENT WHO HAVE FAMILY MEMBER WITH MENTAL DISORDER: A LITERATURE REVIEW

**DOI:** 10.1101/2022.12.09.22282977

**Authors:** Rr Dian Tristiana, Glorino Rumambo Pandin, Ah Yusuf, Moses

## Abstract

Having a family member with mental disorder will affect their healthy family member life. This study to examine the factors impacted on adolescent well-being who have family member with mental disorder. This study was a literature review in two databases SCOPUS and Science which conducted by four steps: (1) identification of literature; (2) screening questions; (3) eligibility using inclusion criteria; and (4) assessment of the quality of the studies. This study found 17 article which then reviewed and analyzed. The study result found seven theme that impacted on well-being included the caregiving responsibilities; the caregiving perception; the caregiving supports; Coping; Caregiving burden; Caregiving positive effects; Psychological impact; Adaptation enhancing. This study the results show the need to explore the seven themes in relation to the conditions of well-being on adolescent who have family member with mental disorder.

## INTRODUCTION

Having a family member with chronic illness will affect their healthy family member life. A study found that one of five children has a parent with mental illness (Reedtz et al., 2018). Children with sibling experiencing mental disorder also have significant adverse effects on the health and well-being of other child (Krzeczkowski et al., 2022). Those children are at high risk for developing mental illness.

Indonesian Basic Health Survey (RISKESDAS) 2018 found that more than 19 million population aged more than 15 experience mental emotional disorders, and more than 12 million people aged over 15 experience depression (Biro Komunikasi dan Pelayanan Masyarakat, 2021). There are various factors which increase the likelihood that children will show disturbance over time. Those factors include being raised in poverty, marital conflict, teen and single parenthood, parental depression, and hostile/angry parenting. Disruption in family structure can lead to several adverse events impacting both the mental health of children and their parents (Behere et al., 2017). Family condition affect the psychological condition in children.

Healthy child development comprises social and emotional wellbeing (some references referred to as non-cognitive skills, subjective wellbeing or character traits), not only simply absence of mental ill-health but the flourishing of positive mental traits (Gregory et al., 2021). Wellbeing and mental health are fundamental rights of children and adolescents vital for sustainable development. Childhood and adolescence are critical developmental periods to foster psychological wellbeing and reduce risk for mental disorders. Disruptions to mental health and wellbeing in childhood and adolescence are correlated with a range of adverse outcomes,

Parent’s mental illness, instead of sensitive, often lead to what has been referred to as avoidant, anxious or insecure/disorganized attachment, which in turn, have been linked to a variety of adverse outcomes (Maybery et al., 2005). Familial stress also has been implicated as a significant predictor of child risk among children of parents with serious mental disorders, even after accounting for parental psychiatric symptomatology and functioning (Tebes et al., 2005). As parental mental health problems are thought to impact negatively on parent-child attachment (Erwin, 1998; Rutter, 1986) it is, therefore, extremely important to examine the factors impacted on adolescent child well-being who have family member with mental disorder.

## METHOD

This review followed the standard Preferred Reporting Items for Systematic Reviews and Meta-Analyses (PRISMA), namely: (1) identification of literature; (2) screening questions; (3) eligibility using inclusion criteria; and (4) assessment of the quality of the studies, which are discussed in the following sections.

### Identification

We searched the literature published until between November 2012 to November 2022 to identify relevant literature, in electronic databases Science Direct, and Scopus, using the following keywords:

| Database | Keyword | Result |
| --- | --- | --- |
| Scopus | (( TITLE-ABS-KEY ( *physical* AND *health* )) OR ( TITLE-ABS-KEY ( *social* AND *well-being* )) OR ( TITLE-ABS-KEY ( *psychological* AND *well-being* )) ) AND ( ( TITLE-ABS-KEY ( *mental* AND *health* )) OR ( TITLE-ABS-KEY ( *mental* AND *disorder* )) OR ( TITLE-ABS-KEY ( *depression* )) OR ( TITLE-ABS-KEY ( *psychiatry* )) ) AND ( ( TITLE-ABS-KEY ( *young* AND *caregiver* )) OR ( TITLE-ABS-KEY ( *young* AND *carer* )) OR ( TITLE-ABS-KEY ( *young* AND *caregiver* )) OR ( TITLE-ABS-KEY ( *caregiving* AND *youth* )) OR ( TITLE-ABS-KEY ( *young* AND *adult* AND *carer* )) OR ( TITLE-ABS-KEY ( *young* AND *adult* AND *caregiver* )) OR ( TITLE-ABS-KEY ( *child* AND *carer* )) OR ( TITLE-ABS-KEY ( *child* AND *caregiver* )) ) ) | 3,102 |
| Science direct | youth carer OR teenager OR adolescent AND well being OR physical OR psychological AND mental illness OR mental disorder OR depression | 39,249 |

### Screening questions

The electronic database search obtained Scopus 3,102 and Science direct 39,249. The double which then screened for the title and abstract, 398 studies were included in the review. Then from the selected articles were screened with the following questions:

1. Were children or teenager or adolescent as carer or caregiver for their parents, sibling or family member with mental illness or mental disorder is included as a general study population? (y/n)
2. Were any kind of social, physical and mental health problems part of the study? (y/n)

Based on the results of the screening questions, 398 studies were first included in the review. Studies were excluded because of limited relevance to children or teenager or adolescent as carers or caregiver for their parents, sibling or family member with mental illness or mental disorder which described the social, physical and psychological aspects of young carers.

### Eligibility using inclusion criteria

The following inclusion criteria were then applied:

Does the study detail the direction on children or teenager or adolescent as carers or caregiver for their parents, sibling or family member with mental illness or mental disorder which described the social, physical and psychological aspects of young carers (+/−/0)?

Among the identified articles, 381 failed to meet the eligibility criteria, hence 17 articles were included in the review (Table 1).

**Table 1.**
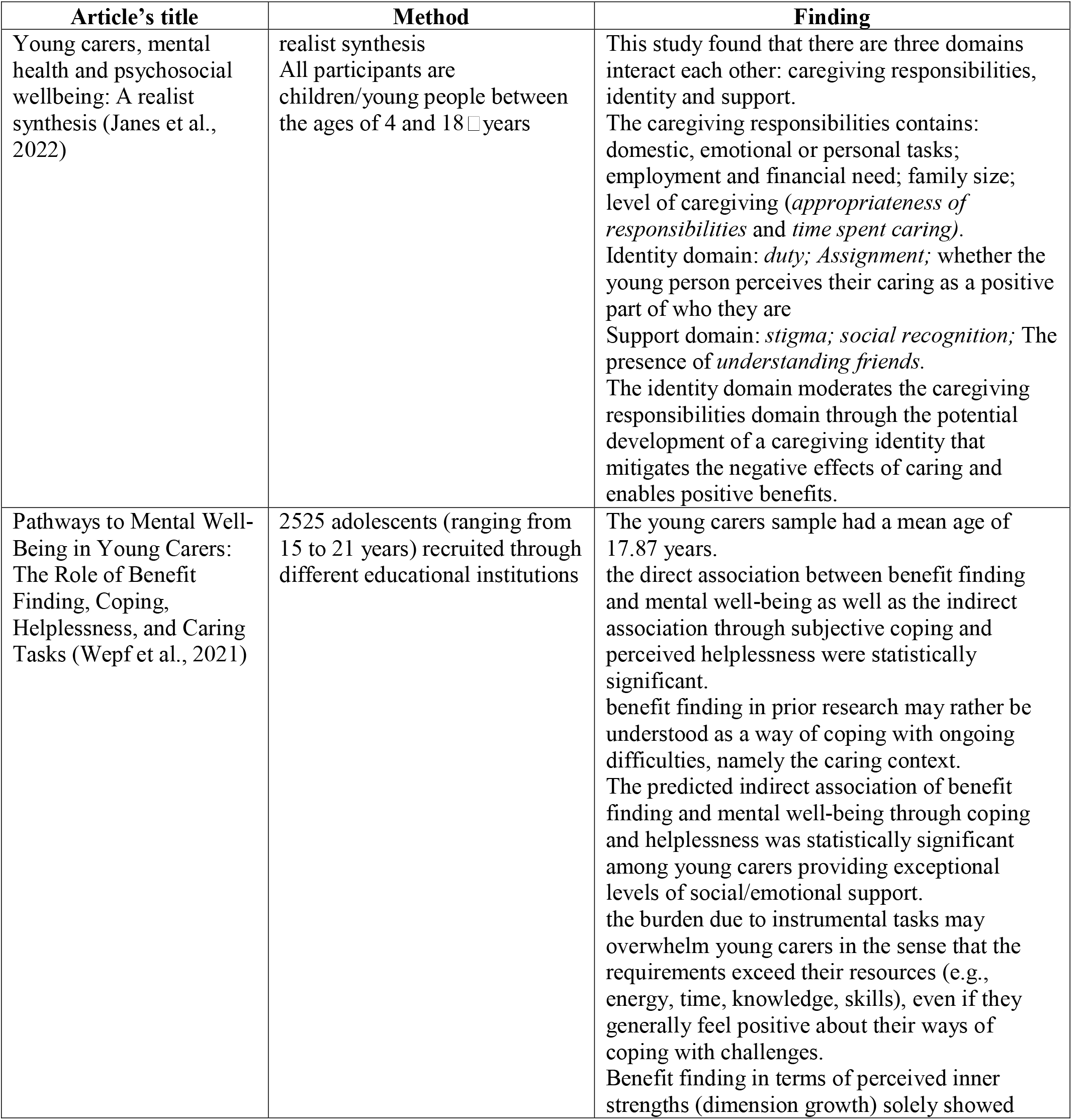
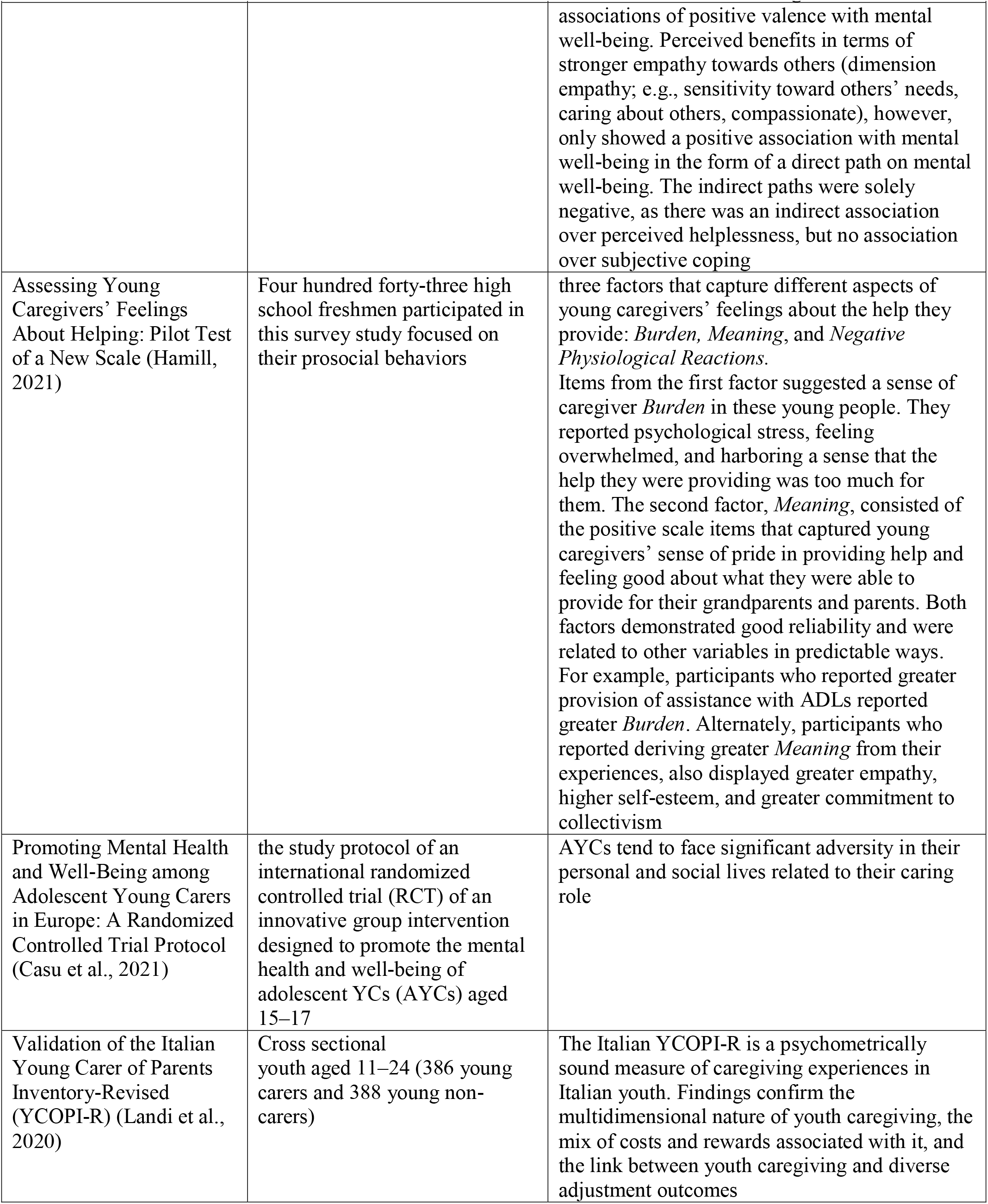
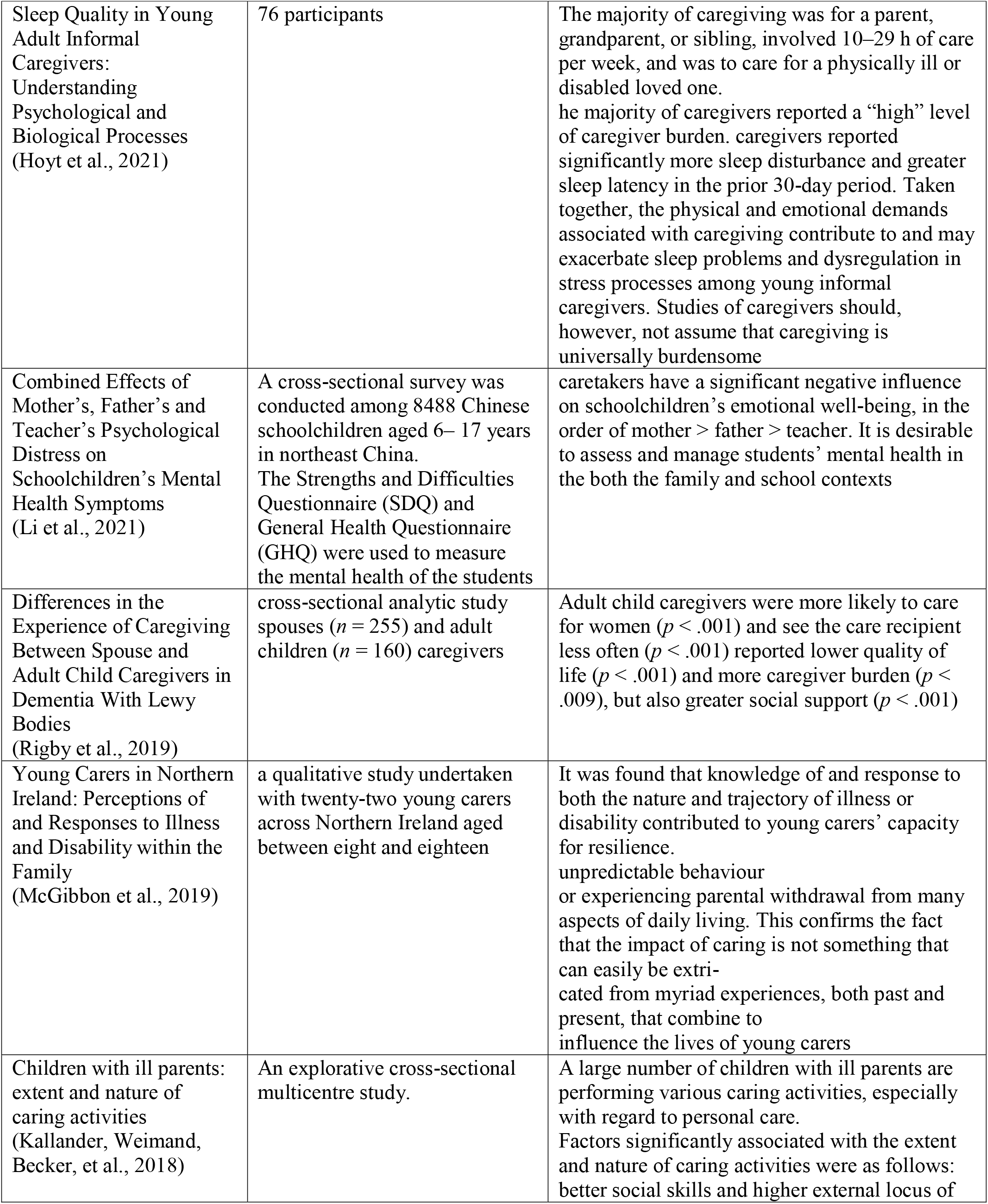
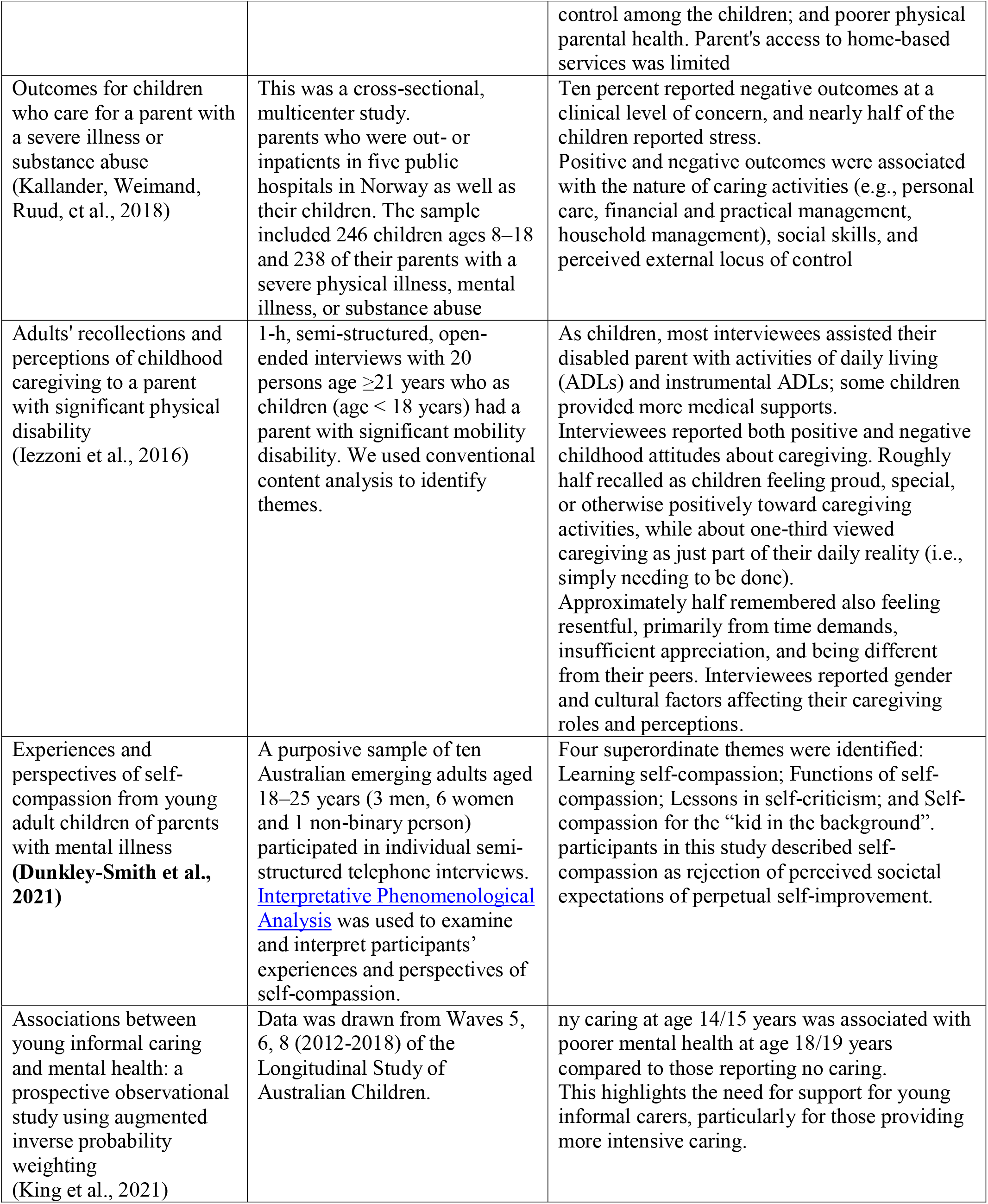
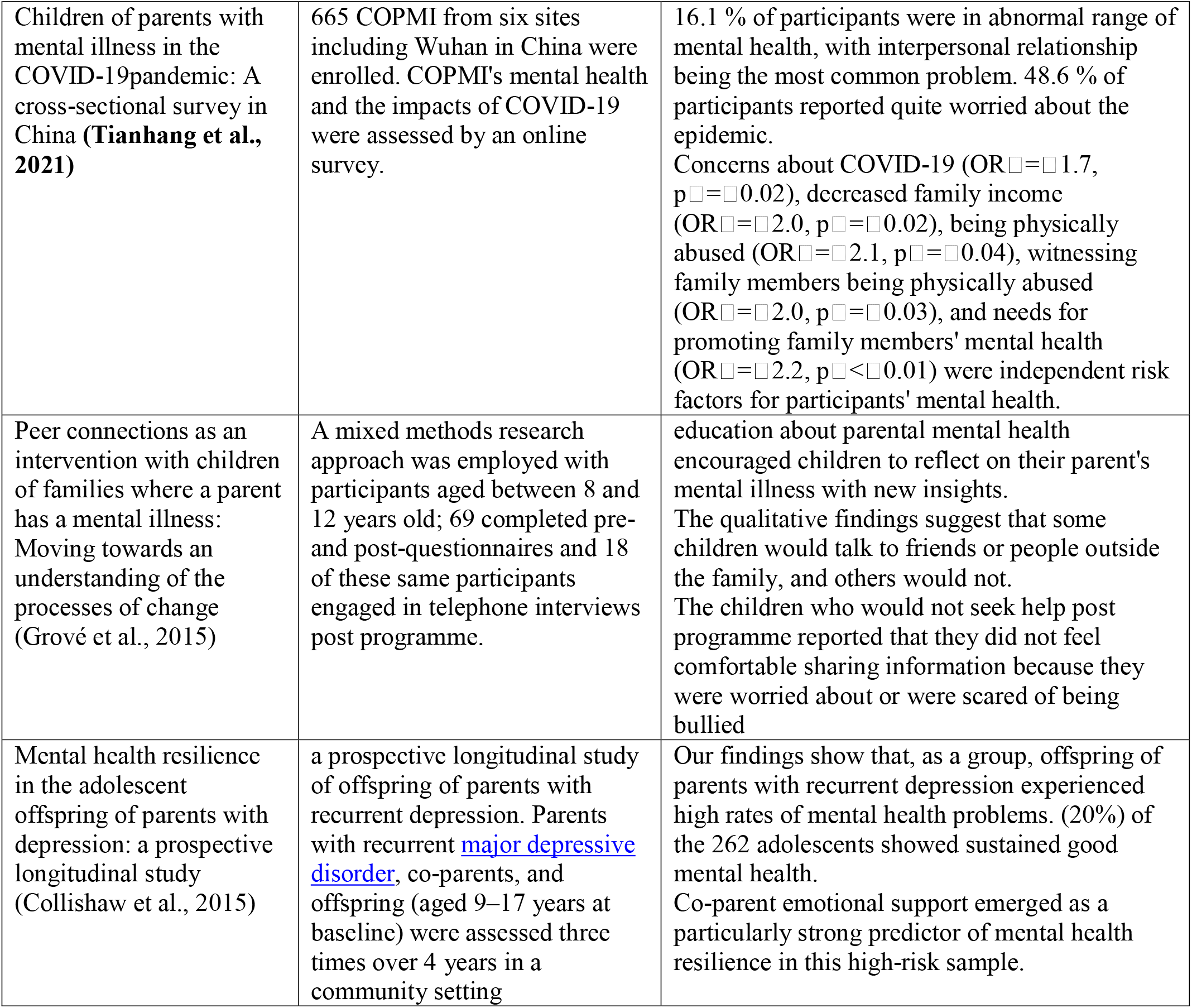
reviewed articles

## RESULT

This review identified within the included studies

### The caregiving responsibilities

This review found that caregiving responsibilities is divided into domestic care or personal tasks (Janes et al., 2022) and caring role (Casu et al., 2021) such personal care (Kallander, Weimand, Becker, et al., 2018) and assisted with activities of daily living (ADLs) and instrumental ADLs (Iezzoni et al., 2016); emotional task (Janes et al., 2022); employment, financial and practical management (Janes et al., 2022; Kallander, Weimand, Becker, et al., 2018), household management and level of caregiving (*appropriateness of responsibilities* and *time spent caring)* (Janes et al., 2022).

### The caregiving perception

The adolescent child views the caregiving activities as identity. This identity is perceived the adolescent child as: *duty; Assignment;* the young person perceives their caring as a positive part of who they are (Janes et al., 2022); perceived helplessness (Wepf et al., 2021) and viewed caregiving as just part of their daily reality (i.e., simply needing to be done) (Iezzoni et al., 2016).

### The caregiving supports

The adolescent child found feeling of support or unsupported from: stigma; social recognition; and the presence of understanding friends (Janes et al., 2022).

### Coping

Subjective coping (Wepf et al., 2021)

### Caregiving burden

The adolescent child harboring a sense that the help they were providing was too much for them (Hamill, 2021); involved 10–29 h of care per week; a “high” level of caregiver burden; sleep disturbance and greater sleep latency in the prior 30-day period (Hoyt et al., 2021), and unpredictable behaviour or experiencing parental withdrawal from many aspects of daily living (McGibbon et al., 2019).

### Caregiving positive effects

Young caregivers’ sense of pride in providing help and feeling good about what they were able to provide for their grandparents and parents. Both factors demonstrated good reliability and were related to other variables in predictable ways. For example, participants who reported greater provision of assistance with ADLs reported greater *Burden*. Alternately, participants who reported deriving greater *Meaning* from their experiences, also displayed greater empathy, higher self-esteem, and greater commitment to collectivism (Hamill, 2021); Roughly half recalled as children feeling proud, special, or otherwise positively toward caregiving activities (Iezzoni et al., 2016); Learning self-compassion; Functions of self-compassion; Lessons in self-criticism; and Self-compassion for the “kid in the background” (Dunkley-Smith et al., 2021) and greater social support (Rigby et al., 2019)

### Psychological impact

The caregiving activities caused a significant negative influence on emotional well-being (Li et al., 2021); psychological stress, feeling overwhelmed, and harboring a sense that the help they were providing was too much for them (Hamill, 2021); lower quality of life; more caregiver burden (Rigby et al., 2019); (Kallander, Weimand, Ruud, et al., 2018); negative outcomes at a clinical level of concern (Iezzoni et al., 2016); feeling resentful, primarily from time demands, insufficient appreciation, and being different from their peers; poorer mental health (King et al., 2021; Tianhang et al., 2021 (Collishaw et al., 2015)).

### Adaptation enhancing

The review found that some condition enhanced adolescent child adaptation that could increase their wellbeing, includes: knowledge and response to both the nature and trajectory of illness or disability (McGibbon et al., 2019); various caring activities, especially with regard to personal care; social skills (Kallander, Weimand, Becker, et al., 2018); perceived external locus of control (Kallander, Weimand, Ruud, et al., 2018); gender and cultural factors (Iezzoni et al., 2016); education about parental mental health (Grové et al., 2015) and Co-parent emotional support (Collishaw et al., 2015).

## DISCUSSION

Caregiving task and responsibilities is related with caregiving demands of caregiver. provide the bulk of assistance with daily activities to adults living with cognitive disabilities. Providing care are seen as the primary stressor, which may trigger secondary stressors such as time conflicts between caregiving and other demands, or strained relationships between caregiver and recipient. This caregiving demand could lead to stress which affect the caregiver well-being.

The caregiving demand is viewed as positive or negative based on the caregiving perception. Identity may be detrimental to well-being when person perceive they are not satisfactorily fulfilling the expectations associated with that identity. Being a caregiver can represent a source of purpose and self-esteem (Rurka et al., 2021). Adult children are able to derive a sense of purpose and meaning from their caregiver identity relies on whether they perceive that they are satisfying the expectations that they have for themselves as a caregiver.

stigma towards mental health in the society is a burden for caregivers when taking care of mentally ill patients (Venkatesh et al., 2016). The caregivers might perceive themselves as being secluded from the society because of limitations on social and leisure activities as well as social stigma and discrimination. perceived social support may be a good predictor of subjective burden (del-Pino-Casado et al., 2018).

Caregiving burden is the level of multifaceted strain perceived by the caregiver from caring for a family member and/or loved one over time (Liu et al., 2020). Prior study found that Burden was affected directly by behavioral problems, frequency of getting a break, self-esteem, and informal hours of care and was not affected by perceived social support (Chappell & Colin Reid, 2002). This study found that caregiving burden involved the hour spent for care, sense providing help, family withdrawal from daily living activity.

Even though caring for family members creates a burden on caregivers, the results of the review also found positive aspects felt by caregivers of adolescents which can reduce the perceived parenting burden. These positive aspects are in the form of pride, meaning giving caregiving, self-compassion and feeling of getting greater social support from others.

The psychological impact felt by caregivers is in the form of decreased emotional well-being, psychological stress, feeling overwhelmed, feeling restful, primarily from time demands, insufficient appreciation, and being different from their peers; poorer mental health.

Several things can encourage adaptation in caregivers, namely knowledge and response to both the nature and trajectory of illness; social skills; perceived external locus of control; gender and cultural factors; education about parental mental health; and Co-parent emotional support.

## Data Availability

All data produced in the present work are contained in the manuscript

## Notes

### Competing Interest Statement

The authors have declared no competing interest.

### Funding Statement

This study did not receive any funding

## REFERENCES

Behere, A. P., Basnet, P., & Campbell, P. (2017). Effects of Family Structure on Mental Health of Children: A Preliminary Study. Indian Journal of Psychological Medicine, 39(4), 457. https://doi.org/10.4103/0253-7176.211767

Biro Komunikasi dan Pelayanan Masyarakat, K. K. R. (2021). Kemenkes Beberkan Masalah Permasalahan Kesehatan Jiwa di Indonesia. Kementerian Kesehatan. https://sehatnegeriku.kemkes.go.id/baca/rilis-media/20211007/1338675/kemenkes-beberkan-masalah-permasalahan-kesehatan-jiwa-di-indonesia/

Casu, G., Hlebec, V., Boccaletti, L., Bolko, I., Manattini, A., & Hanson, E. (2021). Promoting Mental Health and Well-Being among Adolescent Young Carers in Europe: A Randomized Controlled Trial Protocol. International Journal of Environmental Research and Public Health 2021, Vol. 18, Page 2045, 18(4), 2045. https://doi.org/10.3390/IJERPH18042045

Chappell, N. L., & Colin Reid, R. (2002). Burden and well-being among caregivers: Examining the distinction. Gerontologist, 42(6), 772–780. https://doi.org/10.1093/GERONT/42.6.772

Collishaw, S., Hammerton, G., Mahedy, L., Sellers, R., Owen, M. J., Craddock, N., Thapar, A. K., Harold, G. T., Rice, F., & Thapar, A. (2015). Mental health resilience in the adolescent offspring of parents with depression: a prospective longitudinal study. The Lancet Psychiatry, 3(1), 49–57. https://doi.org/https://doi.org/10.1016/S2215-0366(15)00358-2

del-Pino-Casado, R., Frías-Osuna, A., Palomino-Moral, P. A., Ruzafa-Martínez, M., & Ramos-Morcillo, A. J. (2018). Social support and subjective burden in caregivers of adults and older adults: A meta-analysis. PLOS ONE, 13(1), e0189874. https://doi.org/10.1371/JOURNAL.PONE.0189874

Dunkley-Smith, A. J., Reupert, A. E., Ling, M., & Sheen, J. A. (2021). Experiences and perspectives of self-compassion from young adult children of parents with mental illness. Journal of Adolescence, 89, 183–193. https://doi.org/10.1016/J.ADOLESCENCE.2021.05.001

Gregory, T., Sincovich, A., Brushe, M., Finlay-Jones, A., Collier, L. R., Grace, B., Sechague Monroy, N., & Brinkman, S. A. (2021). Basic epidemiology of wellbeing among children and adolescents: A cross-sectional population level study. SSM - Population Health, 15, 2352–8273. https://doi.org/10.1016/J.SSMPH.2021.100907

Grové, C., Reupert, A., & Maybery, D. (2015). Peer connections as an intervention with children of families where a parent has a mental illness: Moving towards an understanding of the processes of change. Children and Youth Services Review, 48, 177–185. https://doi.org/10.1016/J.CHILDYOUTH.2014.12.014

Hamill, S. B. (2021). Assessing Young Caregivers’ Feelings About Helping: Pilot Test of a New Scale. Child and Adolescent Social Work Journal, 38(5), 571–582. https://doi.org/10.1007/S10560-021-00792-7/TABLES/3

Hoyt, M. A., Mazza, M. C., Ahmad, Z., Darabos, K., & Applebaum, A. J. (2021). Sleep Quality in Young Adult Informal Caregivers: Understanding Psychological and Biological Processes. International Journal of Behavioral Medicine, 28(1), 6–13. https://doi.org/10.1007/S12529-019-09842-Y/FIGURES/1

Iezzoni, L. I., Wint, A. J., Kuhlthau, K. A., & Boudreau, A. A. (2016). Adults’ recollections and perceptions of childhood caregiving to a parent with significant physical disability. Disability and Health Journal, 9(2), 208–217. https://doi.org/10.1016/J.DHJO.2015.10.009

Janes, E., Forrester, D., Reed, H., & Melendez-Torres, G. J. (2022). Young carers, mental health and psychosocial wellbeing: A realist synthesis. Child: Care, Health and Development, 48(2), 190–202. https://doi.org/10.1111/CCH.12924

Kallander, E. K., Weimand, B. M., Becker, S., Van Roy, B., Hanssen-Bauer, K., Stavnes, K., Faugli, A., Kufås, E., & Ruud, T. (2018). Children with ill parents: extent and nature of caring activities. Scandinavian Journal of Caring Sciences, 32(2), 793–804. https://doi.org/10.1111/SCS.12510

Kallander, E. K., Weimand, B., Ruud, T., Becker, S., Van Roy, B., & Hanssen-Bauer, K. (2018). Outcomes for children who care for a parent with a severe illness or substance abuse. https://Doi.Org/10.1080/0145935X.2018.1491302, 39(4), 228–249. https://doi.org/10.1080/0145935X.2018.1491302

King, T., Singh, A., & Disney, G. (2021). Associations between young informal caring and mental health: a prospective observational study using augmented inverse probability weighting. The Lancet Regional Health - Western Pacific, 15, 100257. https://doi.org/10.1016/J.LANWPC.2021.100257

Krzeczkowski, J. E., Wade, T. J., Andrade, B. F., Browne, D., Yalcinoz-Ucan, B., Riazi, N. A., Yates, E., Tagalakis, A., & Patte, K. A. (2022). Examining the mental health of siblings of children with a mental disorder: A scoping review protocol. PLOS ONE, 17(9), e0274135. https://doi.org/10.1371/JOURNAL.PONE.0274135

Landi, G., Boccolini, G., Giovagnoli, S., Pakenham, K. I., Grandi, S., & Tossani, E. (2020). Validation of the Italian Young Carer of Parents Inventory-Revised (YCOPI-R). https://Doi.Org/10.1080/09638288.2020.1780478, 44(5), 795–806. https://doi.org/10.1080/09638288.2020.1780478

Li, S., Na, J., Mu, H., Li, Y., Liu, L., Zhang, R., Sun, J., Li, Y., Sun, W., Pan, G., & Yan, L. (2021). <p>Combined Effects of Mother&rsquo;s, Father&rsquo;s and Teacher&rsquo;s Psychological Distress on Schoolchildren&rsquo;s Mental Health Symptoms</p>. Neuropsychiatric Disease and Treatment, 17, 1735–1743. https://doi.org/10.2147/NDT.S302782

Liu, Z., Heffernan, C., & Tan, J. (2020). Caregiver burden: A concept analysis. International Journal of Nursing Sciences, 7(4), 438. https://doi.org/10.1016/J.IJNSS.2020.07.012

Maybery, D., Ling, L., Szakacs, E., & Reupert, A. (2005). Children of a parent with a mental illness: perspectives on need. Australian E-Journal for the Advancement of Mental Health, 4(2), 78–88. https://doi.org/10.5172/JAMH.4.2.78

McGibbon, M., Spratt, T., & Davidson, G. (2019). Young Carers in Northern Ireland: Perceptions of and Responses to Illness and Disability within the Family. The British Journal of Social Work, 49(5), 1162–1179. https://doi.org/10.1093/BJSW/BCY102

Reedtz, C., Lauritzen, C., Stover, Y. V., Freili, J. L., & Rognmo, K. (2018). Identification of Children of Parents With Mental Illness: A Necessity to Provide Relevant Support. Frontiers in Psychiatry, 9(JAN). https://doi.org/10.3389/FPSYT.2018.00728

Rigby, T., Ashwill, R. T., Johnson, D. K., & Galvin, J. E. (2019). Differences in the Experience of Caregiving Between Spouse and Adult Child Caregivers in Dementia With Lewy Bodies. Innovation in Aging, 3(3), 1–15. https://doi.org/10.1093/GERONI/IGZ027

Rurka, M., Jill Suitor, J., & Gilligan, M. (2021). The Caregiver Identity in Context: Consequences of Identity Threat From Siblings. The Journals of Gerontology: Series B, 76(8), 1593–1604. https://doi.org/10.1093/GERONB/GBAA099

Tebes, J. K., Connell, C. M., Ross, E., & Kaufman, J. S. (2005). Convergence of sibling risk among children of parents with serious mental disorders. Journal of Child and Family Studies, 14(1), 29–41. https://doi.org/10.1007/S10826-005-1111-2

Tianhang, Z., Chen, W., Liu, X., Wu, T., Wen, L., Yang, X., Hou, Z., Chen, B., Zhang, T., Zhang, C., Xie, C., Zhou, X., Wang, L., Hua, J., Tang, Q., Zhao, M., Hong, X., Liu, W., Du, C., … Yu, X. (2021). Children of parents with mental illness in the COVID-19pandemic: A cross-sectional survey in China. Asian Journal Psychiatry, 10.1016/j. https://doi.org/10.1016/j.ajp.2021.102801

Venkatesh, B. T., Andrews, T., Parsekar, S. S., Singh, M. M., & Menon, N. (2016). Stigma and mental health- caregivers’ perspective: A qualitative analysis. Clinical Epidemiology and Global Health, 4(1), 23–27. https://doi.org/10.1016/j.cegh.2015.06.003

Wepf, H., Joseph, S., & Leu, A. (2021). Pathways to Mental Well-Being in Young Carers: The Role of Benefit Finding, Coping, Helplessness, and Caring Tasks. Journal of Youth and Adolescence, 50(9), 1911–1924. https://doi.org/10.1007/S10964-021-01478-0/TABLES/6

